# The psychological impact of coronavirus on university students and its socio-economic determinants in Malaysia

**DOI:** 10.1101/2020.10.27.20220723

**Authors:** Muhammad Irfan, Faizah Shahudin, Vincent James Hooper, Waqar Akram, Rosmaiza Binti Abdul Ghani

**Affiliations:** School of Economics and Management, Xiamen University Malaysia; Department of Business Administration, Sukkur Institute of Business Administration Pakistan; Department of Economics, MARA University of Technology

**Keywords:** Anxiety disorder, COVID-19, GAD, Malaysia, Students

## Abstract

This article examines the impact of coronavirus disease 2019 (COVID-19) upon university students’ anxiety level and finds the factors associated with the anxiety disorder in Malaysia. We collected data from 958 students from 16 different universities using an originally designed questionnaire. The Generalized Anxiety Disorder Scale 7-item (GAD-7) was used to estimate the anxiety. We find that 12.3% students were normal, whereas 30.5% were experiencing mild, 31.1% moderate, and 26.1% severe anxiety. Surprisingly, only 37.2% of students were aware of mental health support which was provided by their universities. Moreover, it was found that gender as male (Odds Ratio (OR= 0.798, 95% Confidence Interval (CI)= 0.61 - 1.04)) and having internet access (OR = 0.44, 95% CI= 0.24 - 0.80) were alleviating factors for the anxiety. Whereas, age above than 20 years (OR= 1.30, 95% CI= 0.96 - 1.75), ethnicity Chinese (OR=1.72, 95% CI= 0.95 - 3.1), any other disease (OR=2.0, 95% CI=1.44 - 2.79), decreased family income (OR=1.71, 95% CI=1.34 - 2.17), more time spent on watching COVID-19 related news (OR=1.52, 95% CI=1.17 - 1.97), and infected relative or friends (OR=1.62, 95% CI=1.06 - 2.50) were risk factors for anxiety among students. We suggest that the government of Malaysia should monitor the mental health of the universities’ students more closely and universities should open online mental health support clinics to avoid the adverse impacts of the anxiety disorder.

## 1. Introduction

Coronavirus Disease 2019 (COVID-19) continues to devastate almost every country in the world. As of 24^th^ October 2020, total deaths have reached 1.2 million and infections 42.6 million^1^. The impact of the virus is different in various countries mainly because of the environment, herd immunity, health system, government strategies, and public response. For example, San Marino has maximum per million fatalities (1,237) followed by Peru, Belgium, and Andorra^2^. To avoid the distressing impact of the COVID-19, every country has adopted numerous health, fiscal, and public policies. So far, Malaysia has made tremendous progress in controlling the impact of COVID-19. As a result of the better policies and public responses, as of 11^th^ September 2020, Malaysia has a total 9,628 confirmed cases out of which 9,167 recovered and 128 died (1.33%).

The first case of COVID-19 in Malaysia was confirmed on 25^th^ January 2020 [1]but the cases continued to rise steadily until the end of the February 2020. After the 14^th^ of march the sudden rise in the COVID-19 cases were noted which was believed to be associated with a religious gathering in Kuala Lumpur (Capital city of Malaysia) [2]. Approximately, 16,000 people attended that gathering, eventually leading to a rise in the total cases that were observed during the next two months. The government of Malaysia immediately implemented the Movement Control Order (MCO) commencing from 18^th^ March 2020 to control the spread of the virus [3], which was subsequently relaxed in August 2020. During the period of the MCO gatherings at all places were prohibited including religious services and universities. This was the time when almost all the universities in Malaysia shifted their mode of teaching from physical to virtual.

The sudden change in the mode of teaching due to the potential risk of death caused by COVID-19, isolation, and lockdown have increased the anxiety level and created extreme stress to the general public [4] and students alike [5] The abnormal stress and depression amongst students not only affect their performance but also is associated with heightened self-injury and suicidal attempts [6]. Therefore, it is immensely important to monitor the mental health of the students and assess the risks coupled with preventive factors of the anxiety and associated mental health issues. We selected Malaysia as a case country because of the better possibility of the data collection. Secondly, to date, most of the studies have explored the impact of COVID-19 on students in China, paramedical staff, patients, and even on the general public but the university students in Malaysia are overlooked. Hence, this paper examines the impact ofCOVID-19 on students’ anxiety in Malaysia. As well as it finds the socioeconomic factors which are associated with students’ anxiety. The rest of the paper is organized as follows: Section 1.1 reviews the earlier studies, section 2 explains the data, variables, and methods, section 3 discusses the results, and section 4 concludes the paper.

## 2. Literature Review

A handful of studies attempted to examine the association between COVID-19 and mental health. For instance, Tian et al. (2020) [4] investigated the psychological symptoms of 1,060 Chinese citizens during COVID-19 using a symptoms checklist of 90 via an online questionnaire. They found more than 70% of the total sample had a moderate and higher level of an obsessive compulsion disorder, interpersonal sensitivity, phobic anxiety, and psychoticism. Interestingly, people older than 50 years had relatively more symptoms than young people. Moreover, the symptoms were higher among people who had an education level of undergraduate and below, been divorced, or widowed.

Likewise, Cheng et al. (2020) [7] in Chinese Jiangsu province, measured the impact of COVID-19 on the anxiety level amongst 534 pediatric medical staff. Using a self-rating anxiety scale based upon the Pittsburgh sleep quality index, they found that the prevalence of anxiety was 14.0% amongst the medical staff. They applied stepwise multiple linear regressions to examine the association between anxiety and other socio-economic factors. It was discovered that having a physical condition and concerns relating to the epidemic were positively associated with anxiety levels. Whereas, positional title, better living style, and education were negatively associated with anxiety level. Similarly, another study conducted by Wang et al. (2020) [8] found that there was significantly higher psychological distress amongst medical staff deployed to Hubei province compared to other medical personnel in China.

Most of the studies have only focused on the general public and medical staff (such [as Pieh et al. (2020), Ferrucci et al., (2020), and Huang and Zhao (2020) [9-11]) but ignored college and university students. However, a few studies have attempted to estimate the impact of COVID-19 on the mental health of students. Such as, in China, Cao et al. (2020) [12] examined the psychological impact of the COVID-19 on college students. Using cluster sampling, they collected data from 7,143 students from Changzhi Medical College China. Using GAD-7, they found that 21.3% of respondents were experiencing mild, 2.7% moderate, and 0.9% severe anxiety. Furthermore, they applied the ordered logit model and explored that living in urban areas, living with parents, having a steady family income were protective factors against anxiety. Our study uses the same methods for measuring the anxiety factors associated with this mental health issue. However, our study covers around 16 universities, which produces more generalizable and comprehensive estimates, because of the heterogeneity amongst the sample.

Moreover, Kaparounaki et al. (2020) [13] examined the impact of COVID-19 on the mental health of university students in Greece. They collected the data from 1,000 university students and found that 42.5% of the total sample population had anxiety and 74.3% depression. Using similar data, Patsali et al., (2020) found that depression was present in 12.43% and severe distress was present in 13.46% of the sample population. Of concern, they noticed a 2.59% increase in suicidal thoughts. In Malaysia, we could only find one study by Sundarasen et al., (2020) [14] that was conducted in a similar vein. Their study was the closest to us in terms of our research objectives. However, they used Zung’s self-rating anxiety scale (SAS) to estimate the anxiety level. Quite surprisingly, they found only 8% of students were experiencing mild to moderate and moderate to severe anxiety, which is substantially different from the findings of other studies, hence this requires further investigation. The basic reason for an extremely low number could be using Zung’s self-rating anxiety scale. The anxiety scale has been criticized by earlier studies [15-17] for producing unreliable results. Thus, our study covers a more comprehensive base of 16 universities and applies advanced methods to estimates the anxiety amongst students. We further explored the factors which influence the anxiety level.

## 3. Methods

### 3.1 Data

We created a Google form which consists of 45 questions (including GAD-7) for collecting data and sent the link through email, WhatsApp, Zoom, Microsoft Teams, Google Meetings, WeChat, and other social media platforms. We collected demographic, social, and economic information from the respondents such as living area, age, gender, education, current semester, number of friends, family income, and so on. Whereas, the question related to anxiety was taken from GAD-7. In total, 958 students from 16 different universities participated in the survey. The respondents were requested to provide their consent to use their information for research purposes only. The participation was voluntary and respondents had the option to exit the survey at any point. Data were collected in July 2020 when the mode of teaching in most of the universities was entirely virtual learning.

### 3.2 Variables

The dependent variable “Anxiety Level” was constructed using GAD-7 questionnaire. It is gleaned from the practical self-reporting questionnaire which consisted of 7 items such as feeling nervous, not able to stop worrying, worrying to much about different things, trouble relaxing, being restless, being easily annoyed, and feeling afraid [18]. The GAD-7 is widely used in the literature and its validity has been tested in numerous studies such as Löwe et al., (2008), Barthel et al., (2014), and Seo and Park, (2015) [19-21]. Depending upon the response of the students, four categories of anxiety were created such as 1 means No anxiety, 2 = mild anxiety, 3 = moderate anxiety, and 4 = severe anxiety. Hence, the variable has an order from 1 to 4, where 1 means no anxiety and 4 means severe anxiety. After estimating the anxiety level among students, we applied ordered logit modeling to examine the impact of various socio-economic and demographic variables. The list of our variables of interest along with frequencies and percentages is given in the table 1.

**Table 1.** List of the variables

|  |  |  |  |  |
| --- | --- | --- | --- | --- |
| Anxiety Level (Dependent Variable) | No Anxiety<br>118 (12.3%) | Mild Anxiety<br>292 (30.5%) | Moderate<br>Anxiety<br>298 (31.1%) | Severe<br>Anxiety<br>250 (26.1%) |
| Gender | Male<br>279 (29.1%) | Female<br>679 (70.9%) |  |  |
| Age | Below 20<br>667 (69.6%) | Above 20<br>291 (30.4%) |  |  |
| Ethnicity | Chinese<br>305 (31.8%) | Malay<br>603 (62.9%) | Others<br>50 (5.2%) |  |
| Living Area | Rural<br>286 (29.9%) | Urban<br>672 (70.1%) |  |  |
| Internet Access | Yes<br>917 (95.7%) | No<br>41 (4.3%) |  |  |
| Current semester | Mean<br>2.93 | SD<br>1.61 | Min<br>1 | Max<br>8 |
| Playing sports | Yes<br>655 (68.4%) | No<br>303 (31.6%) |  |  |
| Number of close friends | Mean<br>7.4 | SD<br>5.7 | Min<br>1 | Max<br>28 |
| Living with parents | Yes<br>901 (94.1%) | No<br>57 (5.9%) |  |  |
| Persons living in one home | Mean<br>5.2 | SD<br>1.88 | Min<br>1 | Max<br>13 |
| Personal expenses monthly (RM) | Below 1,000<br>794 (82.9%) | Above 1,000<br>164 (17.1%) |  |  |
| Family income decreased | Yes<br>496 (51.8%) | No<br>462 (48.2%) |  |  |
| Any other disease | Yes<br>156 (16.3%) | No<br>802 (83.7%) |  |  |
| COVID news time spent more than 30 minutes | Yes<br>677 (70.7%) | No<br>281 (29.3%) |  |  |
| COVID infect any relative or friend | Yes<br>83 (8.7%) | No<br>875 (91.3%) |  |  |
| University provides mental health support | No<br>105 (10.9%) | Not sure<br>497 (51.9%) | Yes<br>356 (37.2%) |  |

Author's calculations, SD= standard deviation, RM= Malaysian Ringgit.

**Table 2.** Results of the ordered logit model

| Factors | OR | Std. Error | t value | p-values | C.I 95% |  |
| --- | --- | --- | --- | --- | --- | --- |
|  |  |  |  |  | Lower | Upper |
| Gender (Male) | 0.798* | 0.137 | -1.653 | 0.098 | 0.610 | 1.043 |
| Age above 20 years | 1.297* | 0.153 | 1.694 | 0.090 | 0.960 | 1.752 |
| Ethnicity Chinese | 1.719* | 0.300 | 1.805 | 0.071 | 0.953 | 3.097 |
| Ethnicity Malay | 1.215 | 0.287 | 0.678 | 0.498 | 0.691 | 2.133 |
| Ethnicity Others (reference) |  |  |  |  |  |  |
| Area Urban | 0.831 | 0.132 | -1.406 | 0.160 | 0.641 | 1.076 |
| Internet access (Yes) | 0.446*** | 0.300 | -2.686 | 0.007 | 0.245 | 0.799 |
| Current semester | 0.970 | 0.042 | -0.713 | 0.476 | 0.894 | 1.054 |
| Playing sports (Yes) | 1.053 | 0.132 | 0.390 | 0.697 | 0.812 | 1.365 |
| Number of close friends (log) | 0.972 | 0.077 | -0.366 | 0.715 | 0.837 | 1.130 |
| Living with parents Yes | 0.668 | 0.276 | -1.458 | 0.145 | 0.388 | 1.148 |
| Persons living in one home | 1.042 | 0.033 | 1.247 | 0.212 | 0.977 | 1.112 |
| Expenses above 1,000 RM (yes) | 1.252 | 0.159 | 1.410 | 0.159 | 0.916 | 1.713 |
| Family income decreased (Yes) | 1.706*** | 0.123 | 4.342 | 0.000 | 1.341 | 2.173 |
| Any other diseases (Yes) | 2.001*** | 0.168 | 4.135 | 0.000 | 1.442 | 2.785 |
| COVID news time spent more than 30m | 1.515*** | 0.133 | 3.131 | 0.002 | 1.169 | 1.966 |
| COVID infect any relative or friends | 1.624** | 0.219 | 2.217 | 0.027 | 1.059 | 2.499 |
| Residual deviance | 2467.97 |  |  |  |  |  |
| Akaike information criterion (AIC) | 2505.97 |  |  |  |  |  |
| Observations | 958 |  |  |  |  |  |

Note: authors calculations, OR= Odd Ratios, C.I = Confidence Intervals, and \*p<0.1; \*\*p<0.05; \*\*\*p<0.01

### 3.3 Model

The ordered logit model is suitable when the proportional odds assumption satisfies and the coefficients are usually estimated by using the maximum likelihood method. The model has been used in recent studies by [12,14] to estimate the odds ratios. In our case, we had an ordinal dependent variable “Anxiety level” and our objective was to predict the odds ratios. The odds ratios represent the constant effect of an independent variable on the likelihood that one outcome (dependent variable) will occur.

We had four levels of our dependent variable such as no anxiety, mild anxiety, moderate anxiety, and severe anxiety. The model can be characterized as follows,

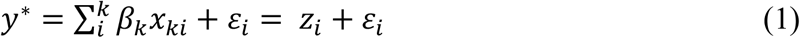

Where *y*^*^ is our unobserved dependent variable and *x*_*ki*_ represents the list of independent variables, *ε*_*i*_ is the error term, and *β*_*k*_ shows the coefficients which we have estimated. Please note that there is no intercept here it means we have estimated m-1 odds ratios that the dependent variable takes on with respect to a particular value identified in equation 1.

## 4. Results and discussion

Table 1 shows the frequencies of anxiety levels among students. Surprisingly, almost 57% of our sampled students were experiencing moderate or severe anxiety during COVID-19, whereas, almost 62% of the students did not know whether their institutions were providing mental health support. Perhaps, universities might not have anticipated the sudden surge in anxiety levels, however, this might have serious repercussions on learning experiences.

The results of the ordered logit model are presented in Table 3. We have only elaborated the significant results where p-value was less than 0.10. We found that the odds of being at the higher-level of anxiety decreases (OR=0.798 95% CI = 0.61 to1.04) if the students were male. In other words, male students were less likely to develop a higher level of anxiety in comparison with female students. Perhaps, female students are more sensitive to the COVID-19 situation than males. Furthermore, the odds of being at the higher-level anxiety increases by 1.3 times (95%CI = 0.96 to 1.75) if the students were above 20 years old in comparison to those whose age was below 20 years. Perhaps older students take the impact of coronavirus more seriously which may lead to an upsurge in anxiety level. Interestingly, the odds of being at a higher level of anxiety increases by 1.7 times (95% CI = 0.95 to 3.09) if the students were of Chinese ethnicity in comparison to other ethnicities (Indian, Bengali, Arabian, and Indonesian).

Moreover, having internet access helps to reduce anxiety amongst students. We found that the odds of being at a higher level of anxiety decreases (OR = 0.45, 95% C. I = 0.25 to 0.80) if students have internet access. Perhaps, the students can have more entertainment options at home which may lead to reduce anxiety. Secondly, due to the lockdown or MCO outdoor sports and other activities were banned, hence, in this situation having internet access could be a significant factor to control anxiety level. Most importantly, students during the MCO period were taking online classes, therefore, having an internet connection helps them to concentrate on the study, which means they would be thinking less of COVID-19.

Undoubtedly, COVID-19 has affected economic activities adversely and because of the lockdown which has led to personal income reducing significantly. This might have a severe impact on the welfare and the anxiety level of individuals. We found that the odds of being at the higher-level of anxiety increase by 1.7 times (95% C.I = 1.34 to 2.17) if students’ family income diminishes because of the coronavirus. Almost 52% of the students reported that their family income has reduced because of the low economic activities and lockdown which was the result of COVID-19.

Furthermore, having any other disease may increase the level of anxiety among students. Our study found that the odds of being at the higher level of anxiety increase by 2.0 times (95% C.I = 1.44 to 2.79) if students have any other pre-existing disease. Perhaps pre-existing diseases have a cumulative effect on fear and lead to a higher level of anxiety. Likewise, spending more than 30 minutes weekly on watching the coronavirus related news can significantly increase the anxiety level among students (OR=1.51, 95% CI = 1.2 to 1.97). Additionally, having an infected friend or relative can significantly increase the anxiety level amongst students. We found that the odds of being at the higher level of anxiety increases by 1.62 times (95% C.I = 1.06 to 2.46) if students’ friends or relatives were infected by COVID-19. The likelihood of having anxiety decreases if the students live with their parents and reside in urban areas. In addition, as the number of close friends increases, anxiety decreases. However, these results were not significant. Perhaps, future larger sample size studies may explore some more interesting exploratory discoveries in our study that may lead to more definite and significant findings. Secondly, the data were collected in July 2020, when the curve of the COVID-19 in Malaysia was getting flatter. Perhaps, data collection in some other time periods may produce slightly different results.

## 5. Conclusion

The COVID-19 outbreak continues to create chaos in almost every country As of September 11^th^ 2020, no vaccination against the virus is available in the market. However, several scientific groups are in the process of developing a potential vaccine. In response to controlling the virus, every country has taken various steps such as lockdowns, quarantines, social distancing, isolations, and MCOs. The fear of death due to COVID-19 and government measures to control the spread of the virus have affected the mental health of citizens and students in particular. The psychological impact of COVID-19 in Malaysian students is a relatively unexplored phenomenon. We explored the anxiety amongst students in Malaysia and found that around 57% of the students were experiencing moderate to severe anxiety. Surprisingly, only 37% of students were aware of mental health support services which are provided by their institutions. Additionally, gender as male and internet access were a protective factor against anxiety, despite the potential for the internet to increase anxiety through constant worrying news reports. Whereas, age above 20 years, ethnicity as Chinese, decreased family income, a pre-existing disease, spending more than 30 minutes weekly on watching or reading COVID-19 related news, and having an infected family or friends were viewed as the major risk factors for anxiety.

Treatment of anxiety involves various interventions including the medication, counselling, and therapy, the combination of all of these usually being more effective. We suggest higher education institutions should assist universities to open mental health support (both psychiatric and psychology based) clinics on every campus, with online access. These services should be provided on both an informal and formal basis as well because those students who are unable to visit the clinic physically can get mental support on call or chat. It is also important to regularly monitor the mental health condition of the students. The financial packages or support can help families to maintain their income which eventually lowers the anxiety level. It can also lead to greater economic productivity in lockdowns. Hence, we suggest the government should also consider providing financial support to the families working from home during lockdowns and higher stringency regimes. Moreover, sports clubs and student societies can also reduce anxiety amongst students by operating within the context of social distancing. These concerns should be addressed by the Ministries of Education and Quality Control. Our policy suggestions can lower the anxiety amongst students and can help the policy setters in formulating the adequate policies. Our results are likely to be of interest to education policy setters around the globe as the majority of students around the world have experienced very similar issues during Covid-19 lockdowns as teaching has been online, and so they confront common issues.

## Data Availability

Selectively available from main author.

## Ethical Statement

We hereby declare that; The paper reflects the authors’ own research and analysis in a truthful and complete manner. The results are appropriately placed in the context of prior and existing research. In addition, no animal or human were harmed, no blood samples were collected, no clinical trials were carried out, identity of the respondents kept confidential, and prior consent from the respondents were taken.

## Footnotes

1 https://www.worldometers.info/coronavirus/

2 https://www.worldometers.info/coronavirus/

## References

[1] R. Pung, C.J. Chiew, B.E. Young, S. Chin, M.I.-C. Chen, H.E. Clapham, A.R. Cook, S. Maurer-Stroh, M.P.H.S. Toh, C. Poh, M. Low, J. Lum, V.T.J. Koh, T.M. Mak, L. Cui, R.V.T.P. Lin, D. Heng, Y.-S. Leo, D.C. Lye, V.J.M. Lee, K. Kam, S. Kalimuddin, S.Y. Tan, J. Loh, K.C. Thoon, S. Vasoo, W.X. Khong, N.-A. Suhaimi, S.J. Chan, E. Zhang, O. Oh, A. Ty, C. Tow, Y.X. Chua, W.L. Chaw, Y. Ng, F. Abdul-Rahman, S. Sahib, Z. Zhao, C. Tang, C. Low, E.H. Goh, G. Lim, Y. Hou, I. Roshan, J. Tan, K. Foo, K. Nandar, L. Kurupatham, P.P. Chan, P. Raj, Y. Lin, Z. Said, A. Lee, C. See, J. Markose, J. Tan, G. Chan, W. See, X. Peh, V. Cai, W.K. Chen, Z. Li, R. Soo, A.L. Chow, W. Wei, A. Farwin, L.W. Ang, Investigation of three clusters of COVID-19 in Singapore: implications for surveillance and response measures, The Lancet. 395 (2020) 1039–1046. https://doi.org/10.1016/S0140-6736(20)30528-6.

[2] K.H.D. Tang, Movement control as an effective measure against Covid-19 spread in Malaysia: an overview, Journal of Public Health. (2020) 1–4. https://doi.org/10.1007/s10389-020-01316-w.

[3] A.Y.A. Bakar, S. Ramli, Psychosocial support for healthcare front liners during COVID-19 pandemic in Malaysia, Asian J. Psychiatry. 54 (2020) 102272. https://doi.org/10.1016/j.ajp.2020.102272.

[4] F. Tian, H. Li, S. Tian, J. Yang, J. Shao, C. Tian, Psychological symptoms of ordinary Chinese citizens based on SCL-90 during the level I emergency response to COVID-19, Psychiatry Res. 288 (2020) 112992. https://doi.org/10.1016/j.psychres.2020.112992.

[5] W. Cao, Z. Fang, G. Hou, M. Han, X. Xu, J. Dong, J. Zheng, The psychological impact of the COVID-19 epidemic on college students in China, Psychiatry Res. 287 (2020) 112934. https://doi.org/10.1016/j.psychres.2020.112934.

[6] M.E. Patsali, D.-P.V. Mousa, E.V.K. Papadopoulou, K.K.K. Papadopoulou, C.K. Kaparounaki, I. Diakogiannis, K.N. Fountoulakis, University students’ changes in mental health status and determinants of behavior during the COVID-19 lockdown in Greece, Psychiatry Res. 292 (2020) 113298. https://doi.org/10.1016/j.psychres.2020.113298.

[7] F.-F. Cheng, S.-H. Zhan, A.-W. Xie, S.-Z. Cai, L. Hui, X.-X. Kong, J.-M. Tian, W.-H. Yan, Anxiety in Chinese pediatric medical staff during the outbreak of Coronavirus Disease 2019: a cross-sectional study, Transl. Pediatr. 9 (2020) 231–236. https://doi.org/10.21037/tp.2020.04.02.

[8] L.-Q. Wang, M. Zhang, G.-M. Liu, S.-Y. Nan, T. Li, L. Xu, Y. Xue, M. Zhang, L. Wang, Y.-D. Qu, F. Liu, Psychological impact of coronavirus disease (2019) (COVID-19) epidemic on medical staff in different posts in China: A multicenter study, J. Psychiatr. Res. 129 (2020) 198–205. https://doi.org/10.1016/j.jpsychires.2020.07.008.

[9] C. Pieh, S. Budimir, T. Probst, The effect of age, gender, income, work, and physical activity on mental health during coronavirus disease (COVID-19) lockdown in Austria, J. Psychosom. Res. 136 (2020) 110186. https://doi.org/10.1016/j.jpsychores.2020.110186.

[10] R. Ferrucci, D. Marino, M.R. Reitano, F. Ruggiero, F. Mameli, B. Poletti, G. Pravettoni, S. Barbieri, A. Priori, A. Averna, Psychological Impact During an Epidemic: Data from Italy’s First Outbreak of COVID-19, Social Science Research Network, Rochester, NY, 2020. https://doi.org/10.2139/ssrn.3576930.

[11] Y. Huang, N. Zhao, Generalized anxiety disorder, depressive symptoms and sleep quality during COVID-19 outbreak in China: a web-based cross-sectional survey, Psychiatry Res. 288 (2020) 112954. https://doi.org/10.1016/j.psychres.2020.112954.

[12] W. Cao, Z. Fang, G. Hou, M. Han, X. Xu, J. Dong, J. Zheng, The psychological impact of the COVID-19 epidemic on college students in China, Psychiatry Res. 287 (2020) 112934. https://doi.org/10.1016/j.psychres.2020.112934.

[13] C.K. Kaparounaki, M.E. Patsali, D.-P.V. Mousa, E.V.K. Papadopoulou, K.K.K. Papadopoulou, K.N. Fountoulakis, University students’ mental health amidst the COVID-19 quarantine in Greece, Psychiatry Res. 290 (2020) 113111. https://doi.org/10.1016/j.psychres.2020.113111.

[14] S. Sundarasen, K. Chinna, K. Kamaludin, M. Nurunnabi, G.M. Baloch, H.B. Khoshaim, S.F.A. Hossain, A. Sukayt, Psychological Impact of COVID-19 and Lockdown among University Students in Malaysia: Implications and Policy Recommendations, Int. J. Environ. Res. Public. Health. 17 (2020) 6206. https://doi.org/10.3390/ijerph17176206.

[15] C.K.W. Schotte, M. Maes, R. Cluydts, P. Cosyns, Effects of affective-semantic mode of item presentation in balanced self-report scales: biased construct validity of the Zung Self-rating Depression Scale, Psychol. Med. 26 (1996) 1161–1168. https://doi.org/10.1017/S0033291700035881.

[16] S. M, B. G, K. M, G. A, T. K, L. M, [Standardization of the Greek version of Zung’s Self-rating Anxiety Scale (SAS)]., Psychiatr. Psychiatr. 23 (2012) 212–220.

[17] T. Yue, Q. Li, R. Wang, Z. Liu, M. Guo, F. Bai, Z. Zhang, W. Wang, Y. Cheng, H. Wang, Comparison of Hospital Anxiety and Depression Scale (HADS) and Zung Self-Rating Anxiety/Depression Scale (SAS/SDS) in Evaluating Anxiety and Depression in Patients with Psoriatic Arthritis, Dermatology. 236 (2020) 170–178. https://doi.org/10.1159/000498848.

[18] R.L. Spitzer, K. Kroenke, J.B.W. Williams, B. Löwe, A Brief Measure for Assessing Generalized Anxiety Disorder: The GAD-7, Arch. Intern. Med. 166 (2006) 1092–1097. https://doi.org/10.1001/archinte.166.10.1092.

[19] B. Löwe, O. Decker, S. Müller, E. Brähler, D. Schellberg, W. Herzog, P.Y. Herzberg, Validation and Standardization of the Generalized Anxiety Disorder Screener (GAD-7) in the General Population, Med. Care. 46 (2008) 266–274.

[20] D. Barthel, C. Barkmann, S. Ehrhardt, C. Bindt, Psychometric properties of the 7-item Generalized Anxiety Disorder scale in antepartum women from Ghana and Côte d’Ivoire, J. Affect. Disord. 169 (2014) 203–211. https://doi.org/10.1016/j.jad.2014.08.004.

[21] J.-G. Seo, S.-P. Park, Validation of the Generalized Anxiety Disorder-7 (GAD-7) and GAD-2 in patients with migraine, J. Headache Pain. 16 (2015) 97. https://doi.org/10.1186/s10194-015-0583-8.

